# Analysing dispatch decision making in high-complexity environments: The P.A.T.H.S. framework

**DOI:** 10.64898/2026.08.14.26360434

**Authors:** Nigel Rees, Jo Angouri, Shawnea Sum Pok Ting, Matthew Booker, Lyba Nadeem, Lauren Williams, David Rawlinson

**Author notes:** Corresponding author: (NR).

## Abstract

**Background:** Decision-making in emergency services involving the allocation of scarce resources is a key challenge for large, complex organisations required to prioritise demand against multiple, often competing criteria. Emergency Medical Dispatch is a case in point, where Enhanced and Critical Care Teams (ECCTs) represent a scarce and lifesaving clinical resource. Despite its operational and system-level significance, the allocation of ECCTs remains under-researched.

**Methods:** We conducted a **methodological development** study using the P.A.T.H.S. framework (Participants, Artefacts, Transition Stages, Historicity, Setting). We designed and piloted this in our previous work under the 999 R.E.S.P.O.N.D. project, which examined the decision-making process for ECCT dispatch. We applied P.A.T.H.S. to 17 dispatch cases (comprising 100 decision-making episodes). We analysed five data sources: recordings of emergency calls and internal dispatch-related interactions, sequence-of-events records, policy documents, and ethnographic observations. A four-phase analysis—indexing & data mapping, transcription & coding, charting, and synthesising & outputs—was undertaken taking Interactional Sociolinguistics as the theoretical approach and methodology.

**Results:** P.A.T.H.S. enables the mapping of non-linear, multifactorial ‘textual trajectories’ across human and non-human actors. The case example presented herein illustrates how information on key risk indicators (e.g. mechanism of injury) were often delayed, fragmented, or lost between the caller, call-handler, and written records. P.A.T.H.S. provides a framework and analytical tool capturing the textual trajectory of information flow, and trace how dispatch decision making unfolds. We subsequently developed a template and codebook for other researchers to further study complex decision making in multi-actoral systems using a textual trajectory approach.

**Conclusion:** This methodological development work demonstrates the potential of P.A.T.H.S. to capture and clarify complex decision-making processes. P.A.T.H.S. offers a practical and theoretically grounded framework for future research, training, and policy that addresses risk points in communication between oral and written forms among teams of actors, to support optimal deployment of scarce resources.

## Introduction

Systems and organisational literature have long shown complex decision making should be understood from a systems perspective [1, 2], recognising that decisions emerge not through individual judgement alone, but through an interaction between human, non-human and technologies, operational and organisational constraints, policy structures, and material conditions. Despite this, there is little research studying interaction in organisation-level decision making by adapting established tools, such as Interactional Sociolinguistics (IS)—the approach we adopt here. This is a gap we are addressing through the study of emergency medical dispatch (EMD). Emergency Medical Services (EMS) worldwide face rising demand within constrained resources, often creating conflicting pressures on already-stretched systems which aim to provide rapid, appropriate, and efficient clinical responses to patients and service users [3–7]. A key area of concern is in the dispatch of Enhanced and Critical Care Teams (ECCTs), which bring specialist expertise to critically ill patients on scene—by Critical Care Practitioners (CCP), doctors, nurses and specialist paramedics, mobilised by Helicopter Emergency Medical Service, rapid response vehicles, or ambulance. ECCTs provide specialist clinical interventions over large geographic areas and are key to improving outcomes for critically ill patients [8, 9]. ECCTs, however, are a scare resource. Despite their significance to patient care, their dispatch has been recognised as an under-characterised link in the response chain; there is little evidence on understanding the whole dispatch decision-making process, not least that could inform deployment optimisation [10–12].

EMD is a dynamic decision-making process shaped by real-time negotiation of risk among multiple actors. It involves the caller and teams of clinical and operational professionals coordinated through a centralised call-handling and resource deployment facility (i.e., a control room like a Clinical Contact Centre or Emergency Operations Centre). Despite an existing (though comparatively small) body of work that has focused on the interaction between the caller and call-handler [13–15], there is scant research bringing interaction analysis tools to a systems approach and studying the entire trajectory of information flow across the possible parts of the response chain [cf. 16].

This results in a lack of evidence about EMD systems [17] and a call for research on the decision-making process in time-sensitive (inter-)actions in high-risk, life-threatening conditions [18]. This paper seeks to contribute to this agenda. It reports the results of a **methodological development** that tests the P.A.T.H.S. framework—a guide to holistically analyse the decision-making process in a multi-actor system—as a novel way of examining ECCT dispatch. We show that P.A.T.H.S and our approach have wide applications across complex organisations.

## Materials and Methods

### Study design

We conducted a methodological development study within the 999 R.E.S.P.O.N.D. programme (RfPPB-21-1847(P)) which researched risk negotiation in dispatch decision making in ECCT in Wales. Emergency calls for the ambulance service in Wales are made via the ‘999’ system. All calls are initially handled through the Medical Priority Dispatch System (MPDS); a scripted and sequentially prompted emergency triage protocol. Call-handlers are required to follow the structure and word choice closely. Emergency calls entering the ‘999’ system are then viewed and interrogated by professionals at the Critical Care Hub—namely a clinician (CCP) and an Allocator—who play a key role in making dispatch decisions for an ECCT.

The primary aim of the present study was to test the usability and value of the P.A.T.H.S. framework (standing for Participants, Artefacts, Transition Stages, Historicity, Setting) for analysing EMD for ECCTs. This work should be understood as a proof-of-concept study: the focus is on **methodological development and accessibility** without necessarily producing generalisable outcomes.

We assembled a multi-modal corpus capturing the EMD workflow and its textual trajectory across the system constituting our focus:

- **Audio recordings of 999 calls** (original calls, and if any, subsequent or duplicated calls, and call-backs to the scene by EMS staff).
- **Audio recordings of internal dispatch-related interactions** (e.g. between dispatchers and other emergency services staff, clinicians, and on-scene responders).
- **Sequence-of-Events (SOE) records from the Computer-Aided Dispatch (CAD) system**, including MPDS outputs, free-text notes, and key event timestamps.
- **Relevant policy and operating procedures** governing ECCT dispatch.
- **Ethnographically-informed observations** of control-room practices.

### Data collection

Data collection was conducted between 1st June 2023 and 23rd December 2024. All audio recordings and SOE records were extracted by the direct clinical care team who routinely access clinical data, from a CAD system (MIS Emergency Systems, Cheshire, UK). Candidate cases were identified from a Structured Query Language database, and listed by date, time, and unique incident ID in spreadsheet form.

All data were redacted and anonymised by the direct clinical care team, then transferred to the research team via secure, encrypted procedures. The research team, including authors of this paper, did not have access to information that could identify individual participations during or after data collection. Audio recordings were anonymised by replacing all spoken identifiable information with an over-recording of a continuous audio tone using Audacity audio processing software. This enabled the team to preserve the original recording timeframe and enabled length of pauses to be retained (important markers of interactional cadence).

A purposive sample of incidents by the direct clinical care team were selected from a list of eligible case numbers, where ECCTs were dispatched during an incident. The cases were divided into core factors which, drawing from our knowledge and frontline experience in EMS and ECC, were likely to show patterns in interactional practices. These include:

- Geographical location of the incident
- Patient age
- Nature of call, such as medical or traumatic
- Times of the day
- Mode of ECC response, such as by air or by road

Cases were eligible for inclusion if:

- The incident had critical care resources dispatched to it at any point in the evolution of the incident for one or more patients of any age.
- A minimum case dataset is available for the incident (including original 999 call recording and SOE.

Cases were excluded if:

- No critical care resources were dispatched at any point.
- The minimum case dataset is not retrievable.
- They resulted in the stand-down of the critical care team before arrival.

As the study was a methodological development study, formal power calculations were not appropriate. Data collection and analysis proceeded iteratively until additional interactions no longer materially altered the analytic account of the interactional patterns relevant to the study’s aims, aligning with Interactional Sociolinguistics (IS) approach (see below) [19].

In total, 17 cases comprising 100 decision-making episodes were analysed, constituting one of the largest corpora of the same kind of studies in the world.

### Ethics

Ethical approvals were obtained (IRAS ID 321594; REC ref: 22/WA/0366). Appropriate data protection impact assessments were completed in collaboration with site-level governance requirements. A Data Protection Impact Assessment was conducted, which helps assess privacy risks to individuals in the collection, use and disclosure of personal information.

#### Analytical approach

Our analysis was informed by **IS** and was concerned with risk negotiation across spoken texts (i.e. speaking) and written texts among different actors in EMD as a complex system.

IS is an established approach in social science to explore patterns of language use in specific settings. It is an ethnographic-oriented, multi-method approach [20, 21] at the interface of anthropology and sociolinguistics. IS is a theory of context through which researchers look into interactions among actors and texts in an organisational-sociocultural context [16]. We examined the data at **micro** (interactional moments within calls), **meso** (team-level decision making and backstage talk), and **institutional** (protocols, Standard Operating Procedures, policy) levels. This enabled us to develop a holistic understanding on how interactants negotiate risk in emergency calls and among EMS professionals through ritualised interactional features. IS enables researchers to effectively translate findings into tailored, system-sensitive training interventions [e.g. 22, 23].

The innovative aspect of this approach lies in combining linguistic micro-analysis with system-level mapping, enabling us to track how information is produced, transformed, and sometimes lost as it moves across different artefacts and actors. This aligns with contemporary sociolinguistic research that emphasises the study of communication within complex professional ecologies, where spoken and written texts co-construct action [21]. Applying this perspective to emergency medical care is novel, offering methodological tools for understanding how teams negotiate risk under pressure and across boundaries of role, medium, and protocol.

The **P.A.T.H.S. framework** [see also 16] provided the organising heuristic to track:

- **Who** acts on information (Participants)
- **What** textual artefacts they use and produce (Artefacts)
- **How** information moves across stages and modalities (Transition Stages)
- **Where** information and artefacts stabilise or change (Historicity)
- **In which setting** decisions occur (Setting).

Visually, P.A.T.H.S. can be represented as per Figure 1.

**Fig 1.**
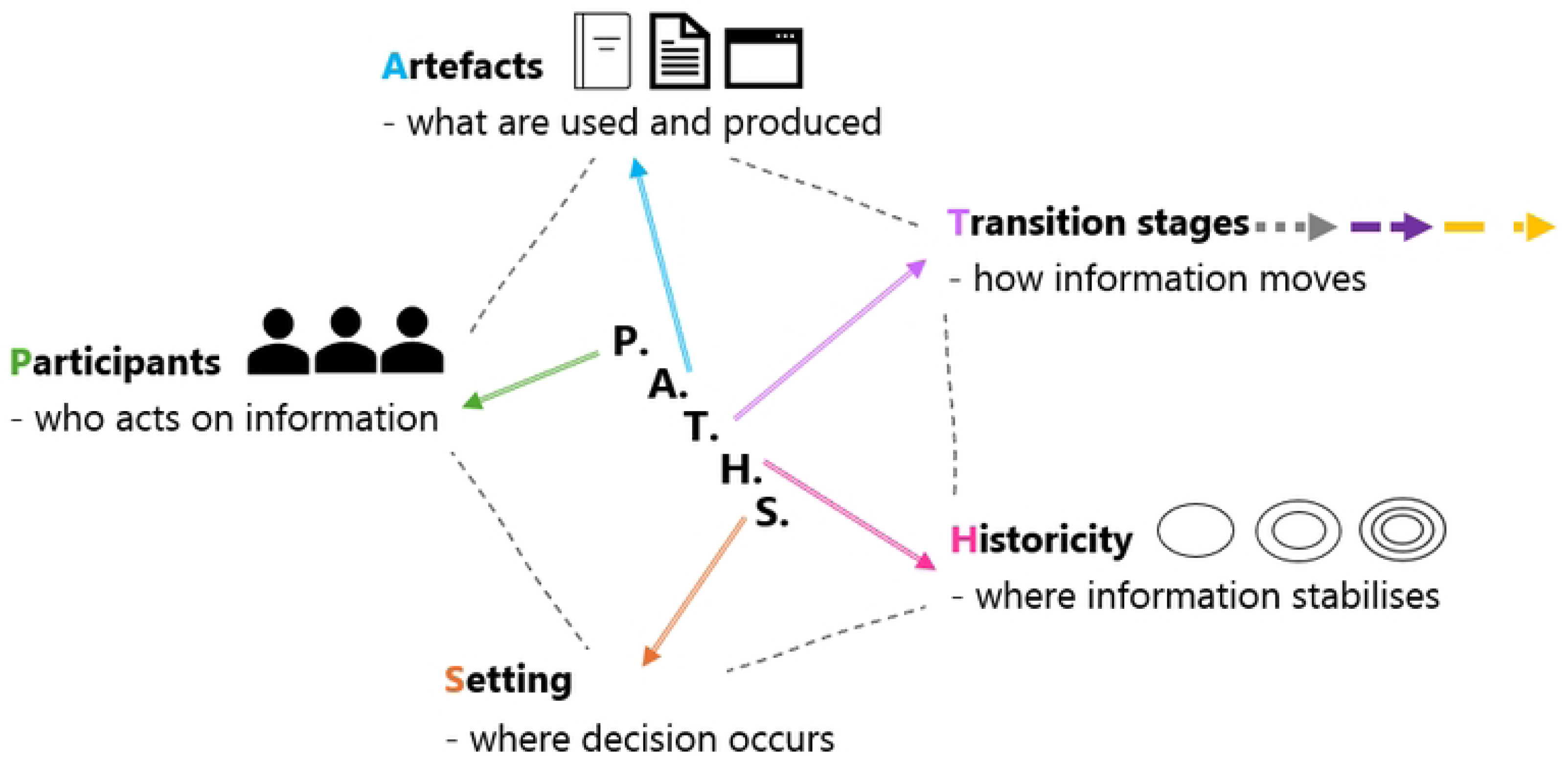
P.A.T.H.S. dimensions.

Identifying and noting these dimensions and their relations enables a mapping of the information landscape, making visible how information, artefacts and actors interact across the decision-making process without imposing a linear sequence. Figure 1 presents a P.A.T.H.S. template (see Figure 6 and 7 for a case-specific visualisation).

Our approach is innovative in general and in EMS research in particular, as it operationalises what are often abstract claims about complexity into a practical, **replicable** method for mapping trajectories of communication and decision-making in complex multi-actor settings. This systematic analysis is structured into four phases, as illustrated below.

### Four phases of analysis of 999 R.E.S.P.O.N.D. using P.A.T.H.S.

#### 1. Indexing and data mapping

We constructed **case-level inventories** linking the core artefacts of each case, i.e. the SOE and related audio recordings (including the original emergency phone call and if any, duplicated calls, call-backs, and internal dispatch-related interactions). The audio recordings were matched to the timestamps on the SOE and its relevant incident ID number. This produced a “data map” per case, which enabled cross-referencing between spoken turns and written artefacts and established a shared timeline for subsequent coding.

#### 2. Transcription and coding

The audio recordings were transcribed using Jeffersonian conventions [24], which captured both the verbal and non-verbal features demonstrated by the call and the call-handler, such as overlaps, pauses, change of pitch and intensity, repetitions, and repair attempts. The transcripts were put in the case-level inventories and “data map”.

We coded the features relevant to risk negotiation, including communication practices that expressed risk and that displayed (a lack of) recognition and uptake of information. We also coded the risk indicators reported by the caller, such as those relating to the mechanism of injury (MOI) and the severity.

Timestamps were created in the transcript of the emergency phone call to match the information or action seen on the SOE, to facilitate the analysis of cross-modality information flow and loss.

In parallel, SOE content was **codified**. This included the Chief Complaint, Reason for Call, patient’s consciousness and breathing, and answers to the scripted Key Questions, input by the call-handler. We also coded MPDS outputs (e.g. the dispatch code and emergency category assigned), free notes left by EMS staff (e.g. to add context, to seek extra information, to give updates on risk, to call for attention, to justify a decision), as well as actions and interventions performed by EMS staff (e.g. to view the case, to allocate or stand down resources).

We then **identified risk indicators** (e.g. cues on MOI, escalation language, caller-call-handler dynamics) across modalities—in (the transcripts of) the audio recordings and in the written SOE—and aligned them to the shared timeline. This prepared for the analysis of information flow, including any acquisition and loss, between textual artefacts and actors.

#### 3. Charting practices

We performed case-level and cross-case-level analyses to examine information flow and loss in individual cases as well as patterns observed across cases.

In the case-level analysis, we juxtaposed the SOE with the transcript of the emergency phone call to compare what was co-constructed by the caller and the call-handler to what was (not) reported on the SOE. This enabled us to identify information transfer and loss when risk indicators were translated from one modality to another, and if applicable, information recovery by other (on-scene) participants later in the SOE. We also read the SOE next to the transcripts of other recordings to see how the information from the SOE might (not) have informed spoken internal dispatch-related interactions (e.g. between a clinician and on-scene responders) which was then selectively reported back to the SOE in writing. Through the side-by-side reading, we tracked the trajectory of risk indicators and the communication practices involved.

For the cross-case-level analysis, we grouped recurrent communication practices (related to, e.g. delayed or fragmented MOI recognition, loss of detail in the SOE relative to the emergency phone call, re-keying by other actors) and charted them against decision points and dispatch outcomes to synthesise the patterns observed. This visualised the link between interactional phenomena and tasking actions or delays.

#### 4. Synthesising and outputs

From the case-level analysis, we created a table for each case to show how risk indicators emerged in the emergency phone call were transferred or lost on the SOE (due to the standardised format of the MPDS protocol) and recovered or added by on-scene responders (with their free narration) (see Table 1 in the next section as example). The table showed the process of risk indicator reduction and expansion, as well as intensification and deterioration of the emergency or the patient’s situation, if applicable.

**Table 1.** Flow and escalation of risk indicators on a case-level analysis.

From the most repeated patterns identified in the cross-case-level analysis, we developed a table synthesising the communication practices that facilitated or impeded information flow and transfer of risk indicators, hence timely and accurate dispatch decision-making (see Table 2 in the next section as example). This table laid the foundation for translating research findings into impact material such as training and policy intervention regarding, for example, points of information loss and question design [e.g. 25].

**Table 2.** Extract of cross-case analysis on communication practices that impacts risk negotiation / transfer of risk indicators.

### A case example

In the following, we show an example to illustrate how we conducted the analysis on the dispatch decision-making process we developed through phase 1-3, supported by our analysis of the context guided by P.A.T.H.S.. We also present the outputs we produced for phase 4. As a representative example, this case—reflective of the rest of our dataset—shows how the P.A.T.H.S.-informed analysis enabled us to see in what ways risk indicators were ‘lost and found’ in the system, thus how dispatch-critical indicators were missed but re-engaged in the decision-making process. We use this case to illustrate how P.A.T.H.S can be applied in other multiprofessional activity systems. We provide a visualisation (see Figure 6 later) building on Figure 1 which seeks to provide a visual template for future studies.

We connected the core artefacts by linking the recording of the emergency phone call—the trigger event in the 999 ecosystem—and the SOE—the anchor record for the decision-making process—in the case-level inventory. We began coding the SOE for what actors in the control room could see from the CAD—the text from which the actors first saw any risk indicators and the platform through which the actors could access the SOE for more information. Here, the Chief Complaint was Traffic/Transportation Accident, the Reason for Call input by the call-handler (a core actor at this stage of the decision-making process) was “fallen off the motorbike –”, the priority level assigned by the MPDS software system (another core actor) was AMBER2 (ranked the lowest among the levels that warrant an emergency response). We then coded other risk indicators on the SOE: the record of the patient’s consciousness and breathing—a yes to both in this case—and the answers to the MPDS Key Questions (Figure 2; timestamps in all Figures represent time passed since the emergency call had started). All these items did not give clear information about the MOI and other risks. It was unclear where the injury occurred and how the accident occurred.

**Fig 2.**
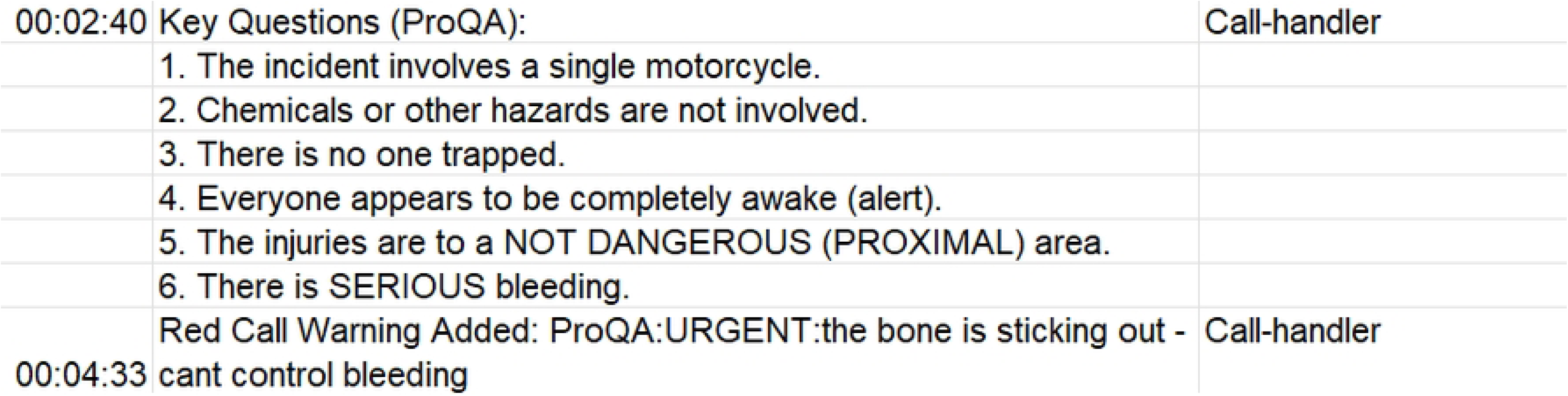
Extract of Sequence of Events record on Key Questions.

Within 2 minutes after the set of answers to the Key Questions appeared, the call-handler added a free note indicating an open fracture and deformity (Figure 2; spelling in all figures and quotes are kept as per original to provide a close representation). Two CCPs (another set of core actors) performed an action—they viewed the case. But they exited the case without any intervention. This suggests the risk indicators visible on the SOE or known to the CCPs as key decision-makers at this stage were inadequate for ECCT dispatch.

The emergency call as the trigger event then ended; the tracing of how information transited from the phone call to the SOE also ended. The caller and call-handler, the core actors in this first stage, moved offstage in our analysis. The SOE, our anchor artefact that would keep evolving, became the focus of our ongoing exploration in the next stage of the decision-making process.

28 minutes after the emergency phone call started, the SOE shows that new actors entered the stage, suggesting possibilities to the decision-making process that we could examine. A dispatcher added a note stating, “police called saying a pt has a compiund fract […] police contact centre unsure if there is still serious bleeding”. The note echoed the previous free note but was not adding new risk indicators, especially regarding the MOI. No action was performed on SOE by the ECCT, indicating the note also had not attracted attention from the key decision-makers. The process had not progressed at that stage. But the SOE displays that there were ongoing interactions unfolding elsewhere (on scene), hinting that we could watch the upcoming entries for potential relevance because information (as resource for decision making) might transit from those sources onto the SOE.

1 hour 31 to 38 minutes after the call started, additional actors came into view. A dispatcher and a second call-handler added five notes relaying information given by police on scene. New risk indicators were added, including one that, for the first time, indicated the injury was at the leg (Figure 3). The second call-handler also added a note calling for actions from other actors—to read these updates, hence learn about these risk indicators. However, not all actors in the control room knew about these additions of risk indicators because (established from our contextual understanding) the CAD would not show that the SOE was updated. No action from the ECCT was recorded on the SOE, suggesting that they might not be aware of the additions and the call for action. But these notes signal that momentum was building in the decision-making process, as information meaningful for decision making had now moved from those interactions into the SOE, the artefact that anchors different actors. This shift marked the need for our close attention to the upcoming entries for a potential breakthrough.

**Fig 3.**
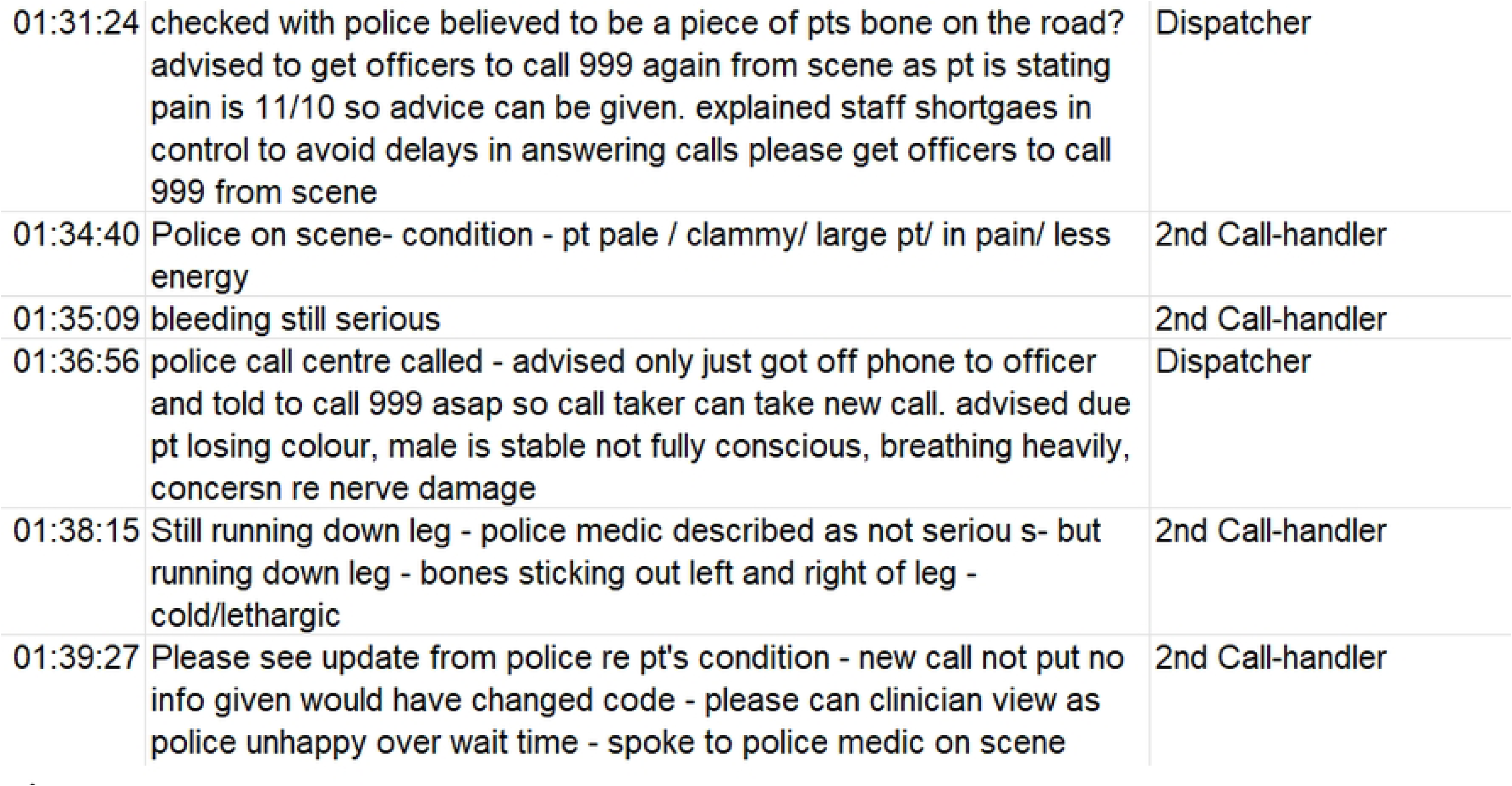
Extract of Sequence of Events record by a dispatcher and a second call-handler.

5 minutes later, multiple entries appear and get intertwined with one another on the SOE. It was a significant moment in the decision-making process where the case got escalated in the system through involvement of multiple additional actors and texts. Among them was the Clinical Support Desk, who first reported that they had viewed the case, signalling that the many risk indicators (eventually) reached the eyes of a clinician. Then they not only requested attention from the ECCT (“Trauma Desk” in Figure 3), but also selected to highlight particular risk indicators that existed on the SOE but were overwhelmed by other information (Figure 4)—amplifying selected indicators and simultaneously supressing other indicators. An Allocator swiftly reported that they had acted on the request. A call-handler supervisor also added that they had asked another centre for more information regarding previous notes left by the second call-handler, which prepared the actors in this control room to expect additional risk indicators. Notes from a duplicated call (from an off-duty paramedic caller) that took place in the meanwhile as another set of resource also entered the SOE (three listed in Figure 4), reiterating risk indicators that appeared before on the SOE but were not highlighted and that informed the MOI.

**Fig 4.**
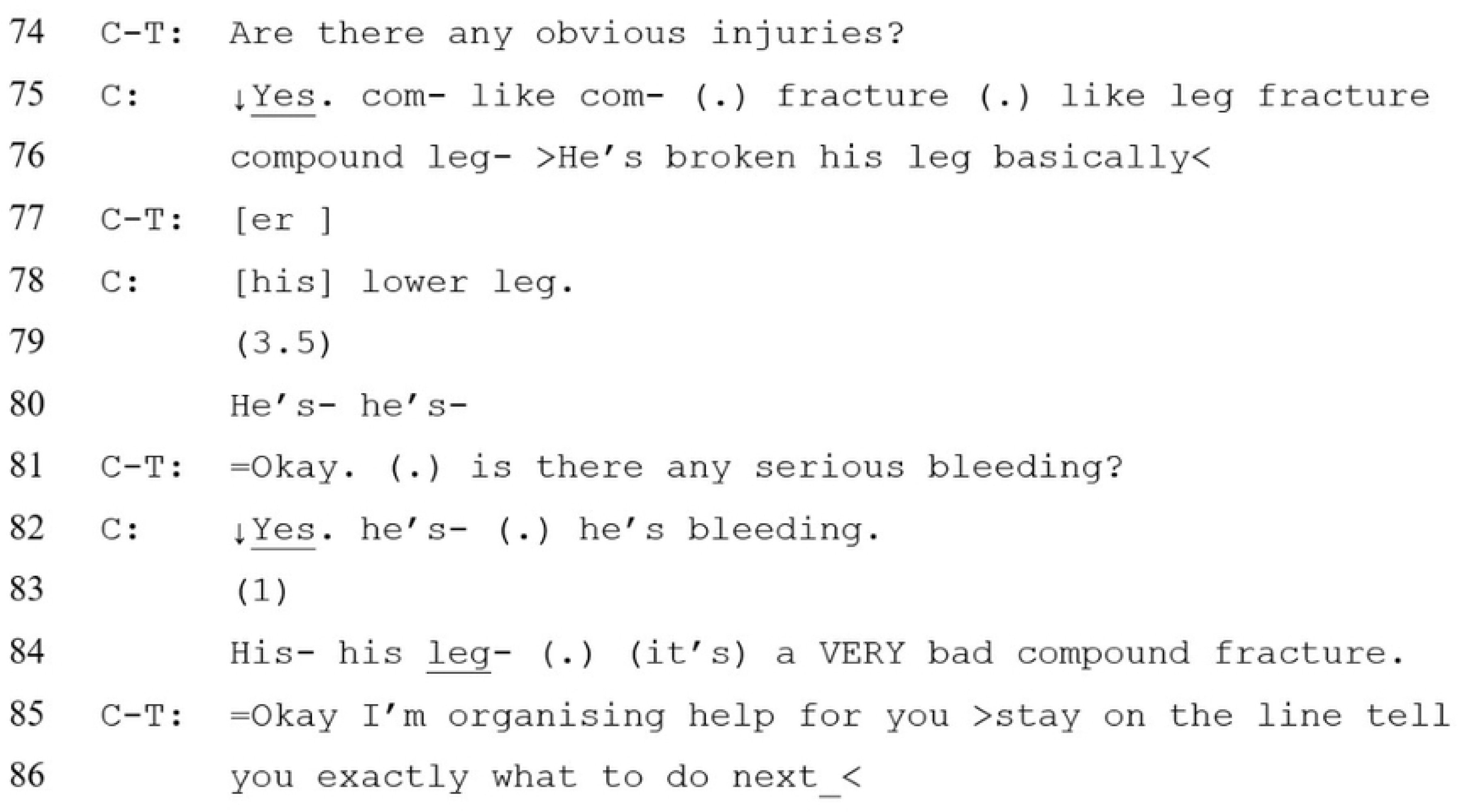
Extract of Sequence of Events record by the Clinical Support Desk and other actors.

The highlighting of selected risk indicators and bringing them to the attention of the ECCT seemed to have contributed to dispatch decision making. 3 minutes later they (“Critical Care Practitioner” in Figure 4) reported that they viewed the case (action one); after another 6 minutes, it is shown on the SOE that they were dispatching a helicopter (action two), which was the first resource allocated to the patient two hours after the call had started.

The coding and analysis of the SOE showed how risk indicators grew then condensed down to inform dispatch decision making. When we read the SOE together with the transcript of the emergency phone call, the two core artefacts in the trigger event in the case-level inventory, we saw how the scripted structure of the MPDS protocol, a significant work system factor in our systems approach, especially its Key Questions, constrained the flow and a timely capturing of risk indicators across modalities—from the caller’s oral report in the emergency phone call to the written SOE. Multiple risk indicators were indeed identified by the caller since the beginning and throughout the call, specifically that the patient had broken his leg. This included when the caller was asked around 2 minutes into the call, “are there any obvious injuries?” (line 74-81 in Figure 5). But the caller’s repsonse was reduced on the SOE to a computer-generated, pre-defined category: “The injuries are to a NOT DANGEROUS (PROXIMAL) area”. A key risk indicator identifed was therefore lost; it was only recovered very late by another actor on scene, the police in this case, on the SOE. A similar information loss occurred with other Key Questions where the injured body parts and other risk indicators reported by the caller were often unrecorded on the SOE (see line 81-86 in Figure 5 as another example).

**Fig 5.**
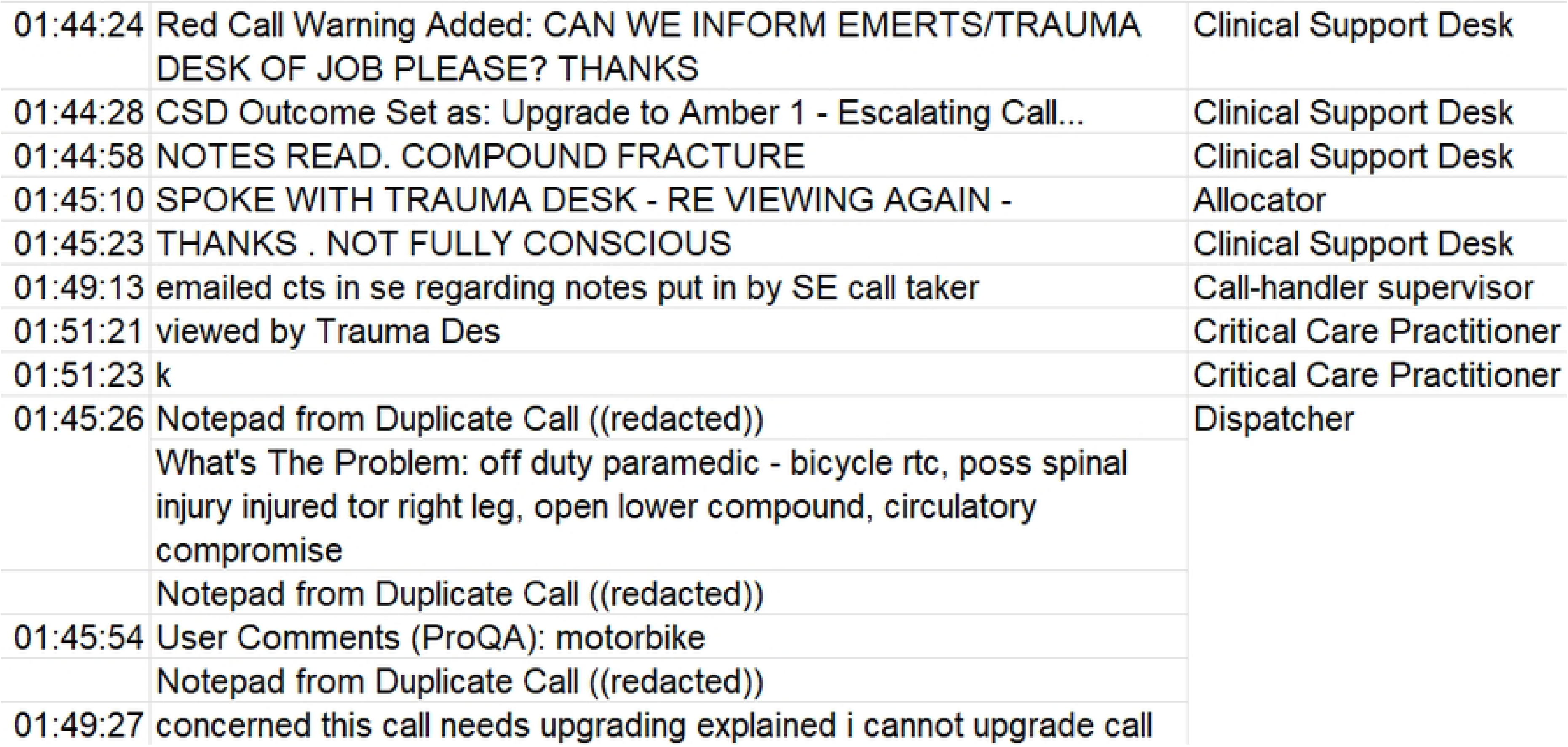
Transcript of the emergency phone call.

**Fig 6.**
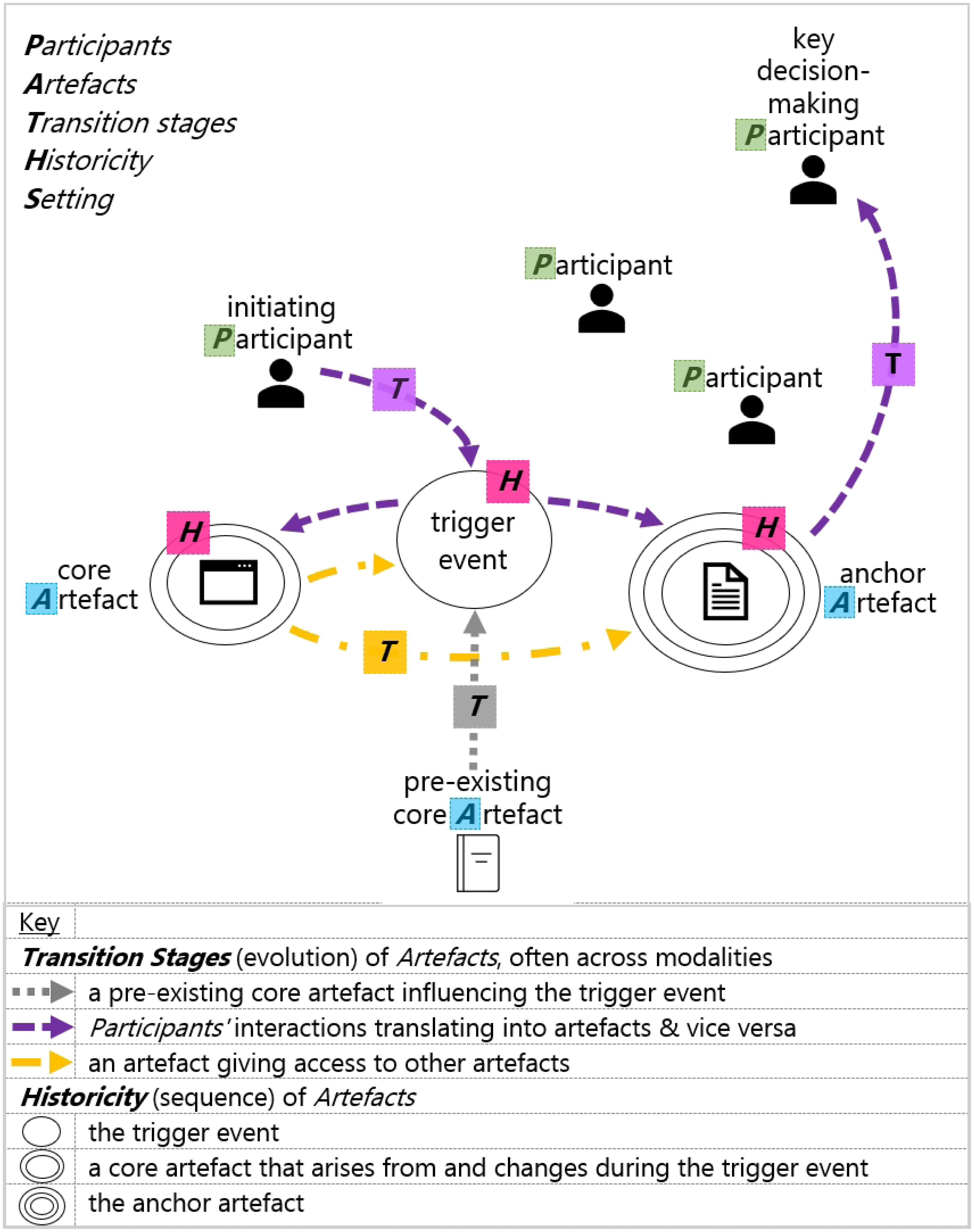
P.A.T.H.S. visual template.

**Fig 7.**
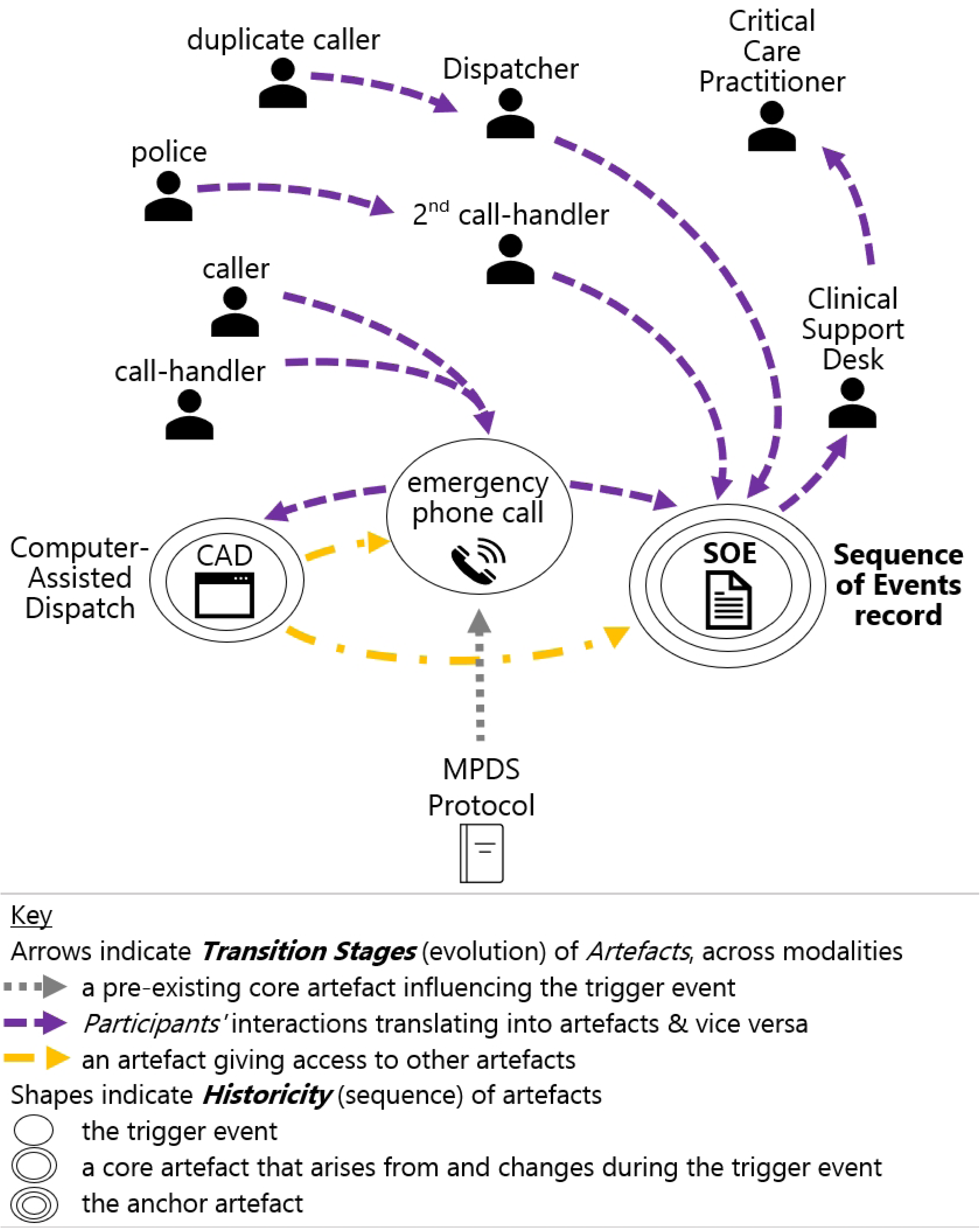
Visual representation of the case example in P.A.T.H.S.

To track and visualise the uptake (or lack of) and flow of risk indicators across the decision-making process starting from the caller in the emergency phone call to the when a CCP made a dispatch decision, we created a table as output entailing the actors appeared in our data map as well as the risk indicators they brought to the case (Table 1). The table enabled us to see that multiple risk indicators were identified in the emergency call. However, they were not translated onto the SOE. The ECCT was therefore not informed in a timely manner about these risks.

Columns from left to right follow the order of appearance of actors; rows from top to bottom follow the first appearance of the risk indicators.

The points of information loss, delay, as well as recovery can be captured and represented holistically through a dynamic visualisation:

From the transcript of the emergency call, we also coded the interaction practices related to risk indication and negotiation—in our work these constituted the mechanisms that explain how and why and when the observed outcome occurs. For example, repetitions and stresses the caller made to emphasise the MOI, reformulations the caller made to articulate the MOI, expansion of the conversational topic (e.g. beyond answering the original question) the caller made to highlight risk, absence of cues from the call-handler to display recognition of risk indicators, verbal cues (e.g. minimal feedback and repetitions) the call-handler made to signal uptake of information, as well as questions the call-handlers posed to seek clarification and confirmation from the caller.

In our cross-case analysis, we compared the patterns of communication practices observed in each case. We created a table as an output to show the most prominent patterns (Table 2 as an extract). The table showed us the way information loss took place in this case example between the emergency phone call and the SOE, which was also observed across other cases. The sequence and design of the scripted protocol—especially with Key Questions—dictated the direction of the narration of the emergency, prioritised and legitimised only certain aspects of risk. The rigid and often-binary design of the computer system, added with time pressure to complete the emergency phone call, made it challenging for call-handlers to pass on the risk indicators reported. The risk captured on the SOE became fragmented and incomplete. Subsequent actors, then, has to mitigate by putting significant work to obtain the information which was, as a matter of fact, offered earlier in the process. This caused delays in dispatch decision making. Also, other parts of the SOE then became important in informing actors for their decision making.

In addition to establishing our understanding on the decision-making process, the communication practices observed in the cross-case analysis also informed our translation of research findings into training materials and interventions to enhance ECCT dispatch [25].

## Results and extra outputs for practice

The analytic approach presented above highlights three areas of significance. First, representative of the patterns observed across the dataset, the case example illustrates how critical information was repeatedly offered by callers yet under-represented in SOE records, delaying dispatch.

Second, the P.A.T.H.S. analysis successfully revealed how ECCT dispatch is shaped by the interplay of:

- **Participants** (call-handlers, dispatchers, clinicians, non-human systems)
- **Artefacts** (emergency calls, SOE, CAD notes, policies)
- **Transition stages** (shifts between verbal and written texts across time)
- **Historicity** (stability vs. change over time)
- **Setting** (the immediate environment).

Third, the case example shows the usefulness and suitabiility of adopting textual trajectories to understand the full story of information flow in the multi-actor decision-making process. Based on our study, we further developed **a template** and a **codebook** to help researchers apply the P.A.T.H.S. model to study complex decision making in multi-actor systems, illustrated here through ECCT dispatch context. The template guides how to capture information flow across cases; the codebook guides an exploration of the transfer, omission, and/or recovery of risk indicators. We present them below.

### Template

The template shows the process and suggested steps for a textual trajectory analysis. The paper draws on ECCT dispatch as an illustration of the process.

**Step (0): Gather contextual factors informed by P.A.T.H.S.**

Before analysis, identify:

- **Participants:** The core actors and their roles, responsibilities, contribution to the decision-making process, locations, and communication modes. (e.g. caller, call-handler, dispatcher, CCP, clinicians, paramedics on scene).
- **Artefacts:** The core texts and their forms/genres (e.g. spoken/audio, written, digital). (e.g. emergency phone call, SOE, MPDS protocol, backstage spoken updates).
- **Transition stages:** How texts progress, move, and evolve across stages. (e.g. audio narrative → written SOE entries → spoken negotiation → later SOE notes).
- **Historicity:** The sequence and start/stop points of texts, any overlap. (e.g. which texts give rise to others, which co-exist, which “freeze” while others continue evolving). Step (1-3): Process of analysis

**Step (1): Identify the trigger event and record interactions in each of its key stage**

Record the following in the emergency call (the spoken/audio text):

- Case entry responses (a yes/no answer for patient’s consciousness and breathing vs expanded caller input).
- Caller’s initial narrative (for the unstructured question “tell me exactly what happened”).
- Responses to the scripted MPDS Key Questions (structured Q&A).
- Free interaction after post-script instructions.
- Communication practices: questioning, confirmation, negotiation.

**Step (2): Trace how information flows into the anchor written record as resource for the decision-making actors**

Identify how the risk indicators marked on the SOE (the structured written text) were transferred or derived from the emergency call(s) (the spoken/audio texts):

- Chief Complaint selected.
- Key Questions + recorded answers (unde the rigid MPDS format).
- Free notes during the live call (e.g. CCP’s input).
- Free notes after call end (e.g. to give contextual updates, justifications).
- Logged actions (allocation of resource, cancellation, relay to other staff).

**Step (3): Explore team interaction and its textual footprint, as mechanisms for decision outcomes**

Follow internal dispatch-related communication as conducted on the SOE and as records translated from spoken interactions (e.g. phone calls):

- Clinical updates (any addition of new risk indicators).
- Dispatch (or non-dispatch) decisions and justifications.
- Action plans proposed.
- Translation of spoken updates back into SOE free notes.
- Communication practices: negotiation between paramedics, CCP, ECC staff.

**Step (4): Synthesise within and across cases as outputs**

Develop case-level mapping of risk indicators with the following:

- **Process focus**:

- Indicators omitted in transfer (e.g. from spoken/audio to SOE).
- Indicators later recovered or re-introduced.
- Indicators multiplied or intensified (e.g. deterioration of patient).
- **Structure**:

- Columns: actors (in chronological order).
- Rows: risk indicators (in order of appearance).

Develop cross-case-level mapping with the following:

- **Process focus**:

- Communication practices that facilitated or inhibited transfer of risk.
- Actions or records observed as outcome.
- **Structure:**

- Columns: communication practices, examples (extracted from transcripts or SOE), outcome observed.

A visual map can be created based on Step 0 to represent the relationship between the dimenisions in P.A.T.H.S.. Figure 7 below is one we created referencing the case example presented.

The visualisation functions both as a product of the analysis and a process for landscaping the decision making trajectory, using P.A.T.H.S. as a framework and tool for studying complex information ecosystems. The work can be further supported by a codebook.

### Codebook

The codebook recommends nine areas that researchers can explore when analysing the core actors and texts identified via P.A.T.H.S.. It informs researchers what to attend to when performing step 1-3 in the template.

1. **Protocol/Structural Constraint**

- Rigid protocol restricts details in an actor’s (e.g. the caller) report.
- *Question (for coding)*: Did the protocol structure itself restrict information recording?
2. **2. Selective Capture of information / Omissions**

- Actors (e.g. call-handlers) recording only part of the narrative.
- *Question*: What was omitted?
3. **3. Summarisation / Compression**

- Long accounts reduced to brief entries.
- *Question*: How much or what narrative detail was lost?
4. **4. Reframing / Re-interpretation**

- Risk indicators being altered in re-telling.
- *Question*: Were severity, urgency, and risk focus shifted?
5. **5. Delay**

- Information given was not recorded immediate but added later (by the same actor).
- *Question*: How did the delay affect the decision-making trajectory?
6. **6. Recovery / Re-introduction**

`o`Previously omitted details re-entered circulation (by other actors).
*Question*: Who re-introduced it, and how was it valued?
7. **7. Amplification / Escalation / Multiplication**

- Risk indicators gained weight as the case escalated.
- *Question*: Did later stages intensify risk framing?
8. **8. Suppression / De-emphasis**

- Risk indicators minimised or sidelined as time passed by.
- *Question*: What was minimised, when, and by whom?
9. **9. Transformation Across Modalities**

- Meaning shifts between audio, written, spoken.
- *Question*: What was lost or changed in relation to the medium?

Together the template and the codebook as tools can support researchers to produce outputs like Table 1 and Table 2 from the case-level and cross-case-level analyses. The outputs visualise the individual processes as well as patterns observed across the datasets, providing solid foundation for further developments of intervention materials. They offer **implementation insights** that looks over (the operation of) the whole system, opening opportunities for training interventions, decision-support tools, and Standard Operating Procedure refinement.

### Feasibility positioning and discussion

#### Feasibility positioning

This study demonstrates the feasibility and innovative potential of the IS-informed P.A.T.H.S. framework in studying complex decision making from a systems perspective, illustrated through emergency medical care’s multi-actor response. It evaluates whether P.A.T.H.S. can be applied consistently, is acceptable and interpretable to varied roles and stakeholders in complex systems—such as clinical and operational professionals in the ECCT dispatch system, and can generate actionable artefacts (templates, codebook, implementation plan). The work establishes a methodological foundation for scaling up to larger datasets, refining the codebook, and embedding P.A.T.H.S. outputs into training and decision-support in complex organisations.

## Discussion

It has long been acknowledged that the evidence base for the accuracy of medical dispatch systems is limited [17]. This is especially true for ECCT dispatch, even though resources are scarce and costly and research has been prioritised for the last decade [10–12]. Increasingly, the value of linguistic analysis of emergency calls is recognised for the contribution in identifying patterns of communication breakdowns as well as the impact of specific word choice and phrasing on subsequent actions and timing, thus dispatch response [26–28]. Alongside this, scholars have called for theoretical and analytic tools capable of examining interrelatedness across written and spoken discourse, including how interpretations are produced by both human and non-human actors within multi-actor environments [29].

Our study demonstrates how the P.A.T.H.S. framework enables researchers to ***reveal and map the decision pathway by following the language***. It shows how each stage of dispatch is shaped by preceding and subsequent actions within the EMD ecosystem. Complex systems are non-linear and adaptive, exhibiting both standardisation and flexibility [30]. Yet the term “complex” has been overused across disciplines as a catch-all for diverse dimensions or simply a substitute for “complicated”. At the same time, complex system research [30, 31] has been expanding, offering conceptual tools for interdisciplinary inquiry [e.g. 32]. This body of work highlights the limitations of analysing decisions as isolated events or discrete interactions. Instead, organisational outcomes emerge through relations between actors, technologies available, institutional and sector-wide procedures, and time-material constraints [cf. 2].

We argue that the EMD of ECCTs constitutes a multi-professional organisational environment in which dispatch decisions are shaped through the alignment of risk perceptions, operational pressures, and resource management across all relevant stakeholders. Existing research demonstrates that dispatch responses are influenced by how the reason for calling and the situation are established, as well as how the problem is indexed and communicated to all the roles involved in the process [33]. These processes extend beyond single encounters and rely upon the coordination of call handlers, dispatchers, clinicians, protocols, and software systems. Organisational research on high-reliability environments similarly demonstrates that decision-making under uncertainty depends upon distributed interpretation [1, 2, 34]. Within EMD, small linguistic shifts may therefore alter pathways and resource allocation decisions across the wider system. Beyond that, P.A.T.H.S. can be operationalised as an analytical and methodological tool for the study of decision trajectories, adding nuance to existing approaches and frameworks.

Our analysis of the textual trajectory surrounding identification of the chief complaint and MOI is particularly significant for ECCT dispatch. Existing research, alongside our findings (see case example), demonstrates that early recognition of the chief complaint and MOI, which may itself be aetiology-dependent, is central to ensuring that ECCTs are dispatched to patients most likely to benefit [9, 35, 36]. Yet the exact aetiology is often unavailable during the initial emergency call. Dispatch decisions therefore depend on provisional classifications produced under time pressure, in this case MPDS codes. Evidence supporting the consistency and accuracy of dispatch systems in identifying acuity nevertheless remains limited [17]. For both clinical and non-clinical professionals involved in call handling and dispatch, interpreting complex situations from verbal information alone presents a substantial challenge. P.A.T.H.S. addresses this by enabling researchers to *reveal the decision pathway by following the language* across different modes and media: tracing how protocol, professional evaluation, and evolving descriptions interact during dispatch decision making.

Using IS as our foundation, we implement P.A.T.H.S. to broaden methodological applications for future work in emergency care response and other complex healthcare systems. We demonstrate how researchers can employ P.A.T.H.S. to track how information and texts evolve as they are circulated, taken up, and linked with one another across a chain of exchanges, as well as to trace how earlier texts influence the way in which later ones are read and interpreted by different actors in the interaction process. It addresses the need for research approaches that both chart the textual trajectory and investigate the elements to which different groups of actors—in line with their roles—orient and respond [e.g. 37].

IS offers analytical value beyond a linguistic analysis of interactions. It has the potential to support multi-professional teams by equipping social and healthcare scientists to collaboratively apply research, which pinpoints where practice breaks down, and translate these findings. IS research has already been informing training and policy interventions developed for frontline medical teams and professionals working in high-stakes healthcare environments [e.g. 22, 23], evidencing its contribution. It offers a blueprint for how interdisciplinary efforts can be realised to tackle complex challenges while maximising expertise, a priority shared by many administrators and policymakers.

### Limitations

We recognise that P.A.T.H.S. has not yet been tested at a larger scale or across multiple sites. We also do not claim that the patterns presented here—identified through the framework—fully capture the diversity of practices across different settings. We would like the reiterate that this exploratory feasibility study is aimed at methodological development and establishing proof-of-concept. Despite scale limitations, this study establishes:

1. The feasibility of using the **IS-informed P.A.T.H.S.** in EMS research and other complex systems.
2. The potential for producing practical outputs (template, codebook, implementation plan).
3. The value of framing dispatch as a **complex socio-linguistic decision-making system**, not a simple technical transaction.

Future work should scale P.A.T.H.S. across larger datasets, refine the codebook, and embed outputs into training and decision support. We hope future research will further advance this work.

## Conclusions

This exploratory feasibility study introduces the P.A.T.H.S. framework as a tool to examine complex decision making from a systems perspective, using ECCT dispatch as an example. P.A.T.H.S. offers a structured and replicable way to unpack complex communication processes, generating templates, a draft codebook, and implementation plan. Further work is needed to refine and extend this approach, but P.A.T.H.S. holds promise for optimising scarce specialist resources and strengthening response systems. By surfacing communication risk points where information is gained or lost—between verbal and written forms and among multiple actors—the framework could help generate much-needed evidence to support optimal deployment.

## Data Availability

All relevant data are within the paper.

## Acknowledgements

We thank all the professionals who participated in our research and generously shared their daily experience, insight and professional practice. We also acknowledge the contribution of our partners at the University of Warwick, University of Bristol, and the Welsh Ambulance Services University NHS Trust for sponsoring the study in collaboration with Emergency Medical Retrieval and Transfer Service and Wales Air Ambulance Charitable Trust.

## Declaration of conflicting interest

The author(s) declared no potential conflicts of interest with respect to the research, authorship, and/or publication of this article.

## Funding statement

The materials presented were collected for the project 999 R.E.S.P.O.N.D. funded by Health and Care Research Wales (RfPPB-21-1847(P)).

## Author contributions

Nigel Rees: Conceptualization, Funding Acquisition, Writing

Jo Angouri: Conceptualization, Data Curation, Formal Analysis, Funding Acquisition, Methodology, Writing

Shawnea Sum Pok Ting: Conceptualization, Data Curation, Formal Analysis, Methodology, Writing

Matthew Booker: Conceptualization, Funding Acquisition, Methodology, Writing – Review & Editing

Lyba Nadeem: Data Curation, Formal Analysis

Lauren Williams: Project Administration

David Rawlinson: Conceptualization, Funding Acquisition, Methodology, Writing – Review & Editing

## Notes

### Competing Interest Statement

I have read the journal's policy and the authors of this manuscript have the following competing interests: NR Receives funding from HCRW and NIHR and is an employee of WAST All authors are now a in receipt of funding from NIHR

### Clinical Trial

NA

### Author Declarations

Ethical approvals were obtained (IRAS ID 321594 REC ref: 22/WA/0366). Appropriate data protection impact assessments were completed in collaboration with site-level governance requirements. A Data Protection Impact Assessment was conducted, which helps assess privacy risks to individuals in the collection, use and disclosure of personal information

